# Early risk assessment for COVID-19 patients from emergency department data using machine learning

**DOI:** 10.1101/2020.05.19.20086488

**Authors:** Frank S. Heldt, Marcela P. Vizcaychipi, Sophie Peacock, Mattia Cinelli, Lachlan McLachlan, Fernando Andreotti, Stojan Jovanović, Robert Dürichen, Nadezda Lipunova, Robert A. Fletcher, Anne Hancock, Alex McCarthy, Richard A. Pointon, Alexander Brown, James Eaton, Roberto Liddi, Lucy Mackillop, Lionel Tarassenko, Rabia T. Khan

**Author notes:** Corresponding author: Correspondence address: Rabia Tahir Khan, Sensyne Health plc, Schrodinger Building, Heatley Road, Oxford Science Park, Oxford, OX4 4GE.

## Abstract

**Background:** Since its emergence in late 2019, the severe acute respiratory syndrome coronavirus 2 (SARS-CoV-2) has caused a pandemic, with more than 4.8 million reported cases and 310 000 deaths worldwide. While epidemiological and clinical characteristics of COVID-19 have been reported, risk factors underlying the transition from mild to severe disease among patients remain poorly understood.

**Methods:** In this retrospective study, we analysed data of 820 confirmed COVID-19 positive patients admitted to a two-site NHS Trust hospital in London, England, between January 1^st^ and April 23^rd^, 2020, with a majority of cases occurring in March and April. We extracted anonymised demographic data, physiological clinical variables and laboratory results from electronic healthcare records (EHR) and applied multivariate logistic regression, random forest and extreme gradient boosted trees. To evaluate the potential for early risk assessment, we used data available during patients’ initial presentation at the emergency department (ED) to predict deterioration to one of three clinical endpoints in the remainder of the hospital stay: A) admission to intensive care, B) need for mechanical ventilation and C) mortality. Based on the trained models, we extracted the most informative clinical features in determining these patient trajectories.

**Results:** Considering our inclusion criteria, we have identified 126 of 820 (15%) patients that required intensive care, 62 of 808 (8%) patients needing mechanical ventilation, and 170 of 630 (27%) cases of in-hospital mortality. Our models learned successfully from early clinical data and predicted clinical endpoints with high accuracy, the best model achieving AUC-ROC scores of 0.75 to 0.83 (F1 scores of 0.41 to 0.56). Younger patient age was associated with an increased risk of receiving intensive care and ventilation, but lower risk of mortality. Clinical indicators of a patient’s oxygen supply and selected laboratory results were most predictive of COVID-19 patient trajectories.

**Conclusion:** Among COVID-19 patients machine learning can aid in the early identification of those with a poor prognosis, using EHR data collected during a patient’s first presentation at ED. Patient age and measures of oxygenation status during ED stay are primary indicators of poor patient outcomes.

## Introduction

COVID-19, caused by the severe acute respiratory syndrome coronavirus 2 (SARS-CoV-2), is a novel infectious disease that leads to severe acute respiratory distress in humans. In March 2020, the World Health Organisation declared the outbreak a pandemic and, by May 19^th^, it had caused more than 4 800 000 confirmed cases and 310 000 deaths worldwide [1]. Disease severity for COVID-19 appears to vary dramatically between patients, including asymptomatic infection, mild upper respiratory tract illness and severe viral pneumonia with acute respiratory distress, respiratory failure and thromboembolic events that can lead to death [2–4]. Initial reports suggest that 6%-10% of infected patients are likely to become critically ill, most of whom will require mechanical ventilation and intensive care [3,5].

Currently, few prognostic markers exist to forecast whether a COVID-19 patient may deteriorate to a critical condition and require intensive care. In general, patients can be grouped into three phenotypes, being at risk of thromboembolic disease, respiratory deterioration and cytokine storm [6]. Early clinical reports find that age, sex and underlying comorbidities, such as hypertension, cardiovascular disease and diabetes, can adversely affect patient outcomes [7,8]. However, few studies have leveraged machine learning to systematically explore risk factors for poor prognosis.

Increasingly, hospitals collate large amounts of patient data as electronic healthcare records (EHRs). Combined with state-of-the-art machine learning algorithms, these data can help to predict patient outcomes with greater accuracy than traditional methods [9,10]. However, EHR data for COVID-19 remains scarce in the public domain, prompting many authors to focus on statistical analyses instead [11–14]. Where machine learning has been applied to COVID-19, results have been promising, but most studies suffer from a lack of statistical power owing to small sample size [15–18]. Jiang *et al*. applied predictive analytics to data from two hospitals in Wenzhou, China, which included 53 hospitalised COVID-19 patients, to predict risk factors for acute respiratory distress syndrome (ARDS) [15]. Exploring the risk factors for in-hospital deaths, Zhou and co-workers used univariate and multivariate logistic regression on data of 191 patients in two hospitals in Wuhan, China [16]. Similarly, Xie *et al*. used logistic regression to predict mortality, training a model on 299 patients and validating it on 145 patients from a different hospital in Wuhan, China [18]. Gong *et al*. used a logistic regression model to identify patients at risk of deterioration to severe COVID-19, applied to the data of 189 patients in Wuhan and Guangdong, China [17].

A key factor that determines the success of risk prediction models is the quality and richness of the available data. Studies to date have used a combination of demographics, comorbidities, symptoms, and laboratory tests [15–17,19]. These data typically comprise the patients’ entire historical record, as well as observations collected during the current hospital stay [16,18–20]. While the inclusion of a patient’s full EHR history improves predictive performance, such approaches may be limited in their clinical applicability to early risk-assessment; at the point of presentation in hospital, the entire EHR of a patient is rarely available.

In this work, we retrospectively apply machine learning to data of 820 confirmed COVID-19 patients from two tertiary referral urban hospitals in London to predict patients’ risk of deterioration to one of three clinical endpoints: A) admission to an adult intensive care unit (AICU), B) need for mechanical ventilation, and C) in-hospital mortality. We restrict our analysis to EHR data available during a patient’s first presentation in the emergency department (ED) as this more accurately resembles the hospital reality of early-risk assessment and patient-stratification. Our analysis provides a proof of principle for COVID-19 risk assessment, with models achieving a high prediction performance, indicating that patient age, oxygenation status and selected laboratory tests are prime indicators of patient outcome.

## Methods

### Data collection and study design

Anonymised EHR data of patients admitted to two hospitals in London, England, between January 1^st^, 2020 and April 23^rd^, 2020, were gathered by Chelsea & Westminster NHS Foundation Trust (NHS Trust, hereafter). The data was supplied in accordance with internal information governance review, NHS Trust information governance approval, and General Data Protection Regulation (GDPR) procedures outlined under the Strategic Research Agreement (SRA) and relative Data Sharing Agreements (DSAs) signed by the NHS Trust and Sensyne Health plc on 25th July 2018.

Data encompasses clinical observations collated from inpatient encounters. The analysis was restricted to adult patients aged between 18 and 100 years at the time of their most recent hospital admission (assumed to be the COVID-19-related admission). Only confirmed SARS-CoV-2 positive patients, as determined by quantitative reverse-transcription PCR (qRT-PCR), were included. 65% of patients were male and 35% female (Table 1). The majority were white British (28%) or did not state their ethnicity (24%) (see also Fig. S1). All clinical features and their coverage in the data set are listed in Table S1. Features include patient demographics (3 in total), vital signs (4 in total), laboratory measurements and clinical observations (60 in total). For vital signs and laboratory measurements, patients may have received multiple test results during their stay. These values were aggregated for each feature to only retain the respective minimum, maximum, mean and last observation value. Only clinical features with at least 5% coverage in the patient population were considered. The data set covered the patient’s entire encounter history from their admission to the hospital’s ED, with a median length of stay in that department of 5 hours, to their discharge. The median length of in-hospital stay was 7.2 days.

**Table 1.** Composition of patient population.

| Demographics |  |
| --- | --- |
| Patient age (years) |  |
| Range | 18-100 |
| Overall mean (standard deviation) | 67.3 (16.8) |
| Female mean (standard deviation) | 70.3 (17.2) |
| Male mean (standard deviation) | 65.8 (16.4) |
| Sex (number of patients) |  |
| Female | 286 (34.9%) |
| Male | 533 (65.0%) |
| unknown | 1 (0.1%) |
| Ethnicity (number of patients) |  |
| White British | 230 (28%) |
| Not Stated | 196 (23.9%) |
| Ethnic Other | 97 (11.8%) |
| White Other | 76 (9.3%) |
| Asian Indian | 63 (7.7%) |
| Asian Other | 39 (4.8%) |
| Unknown | 29 (3.5%) |
| Black African | 24 (2.9%) |
| Black Caribbean | 23 (2.8%) |
| Asian Pakistani | 11 (1.3%) |
| Black Other | 10 (1.2%) |
| Others | 22 (2.7%) |

### Cohort definition

A total of 3229 patients fell within the observation time and study parameters. From these patients, three cohorts were derived, one for each clinical endpoint, as follows (see Fig. S2 for flow diagram and patient numbers). Only confirmed COVID-19 positive patients were considered. Patients who did not have information relating to an admission to any hospital department in 2020 were excluded. Furthermore, the following exclusion criteria were applied to each of the considered endpoints: for cohort A) patients without a documented ward location were excluded; for cohort B) patients without information on oxygen supply were excluded; for cohort C) patients without hospital discharge information were excluded. Finally, since our models were trained on data available during a patient’s stay in the ED, we removed patients who did not have a documented ED visit.

Each cohort was divided into target and control groups (see Table 2). For AICU admission, target patients comprise those that were admitted to an AICU at any time during their hospital stay, while control patients are those that remained in any other ward for their entire admission. Target patients in the ventilation cohort were defined as requiring invasive mechanical ventilation, whereas control patient required no or only minimal breathing assistance. Both categories are based on clinical records of oxygen supply according to Table 3. Note that from clinical data the total number of mechanically ventilated patients was 135, however only 62 were visible in our data. This results from staggered deployment of EHR data in the two hospitals such that one site is understood to lack certain data related to mechanical ventilation. Mortality data was based on the discharge destination (mortuary) in clinical records. All regularly discharged patients or patients remaining in hospital were considered alive.

**Table 2.** Clinical endpoint cohorts.

|  | Cohort A<br>(AICU admission) | Cohort B<br>(ventilation) | Cohort C<br>(mortality) |
| --- | --- | --- | --- |
| Number of patients | 820 | 808 | 630 |
| Target patients | 126 (15%) | 62 (8%) | 170 (27%) |
| Control patients | 694 (85%) | 742 (92%) | 460 (73%) |

**Table 3.** Target and control definition for ventilation cohort.

| Category | Clinical observation value |
| --- | --- |
| Control | room air, air/none, nasal cannulae, high flow nasal cannulae, venturi mask, face mask, non-rebreather mask, simple face mask, swedish nose with, oxygen, mask, HFOV, face/tracheostomy mask, CPAP, BiPAP |
| Target | ventilator, tracheostomy, CMV, VC-CMV, t-piece, HELIOX, IPPV, SIMV, PC-BIPAP, APRV, CPAP / ASB_SPN / CPAP/PS |

### Patient outcome prediction

Three machine-learning algorithms were benchmarked to predict patient outcomes from EHR data: logistic regression, random forest and Extreme Gradient Boosted Trees (XGBoost). Logistic regression, which predicts the probability of a clinical endpoint as a linear function of the feature space, was used as a baseline algorithm. The model was regularised with elastic net using equal weighting given to L_1_ and L_2_ penalties in order to account for the high dimensionality of the data set relative to the number of observations. A random forest [21], i. e., an ensemble of decision trees where each tree is trained on a slightly different subset of data, was trained using 100 trees and splits were evaluated using Gini impurity. Classes were inversely weighted to account for the class imbalance present in the data set. An XGBoost algorithm [22] was trained with its hyperparameters set to 100 trees, max tree-depth of 6, step-shrinkage of 0.3, no subsampling and L_2_ regularisation, to minimize log-loss. This tree-based algorithm trains decision trees sequentially, with each new tree being trained on the residuals of previous trees.

### Performance evaluation

All models were evaluated using a stratified 3-fold cross-validation strategy. Results are reported as mean and standard deviation across these folds. Predictive performance was measured in terms of area under curve (AUC) of the receiver operating characteristic (ROC) as well as F1 score at each model’s ideal classification threshold as derived from the ROC curve. Given the presence of class-imbalance, precision-recall curves were also computed to assess expected real-world performance relative to random classifiers.

In order to extract the clinical features most relevant to predictions, permutation feature importance (PFI) was calculated for each model post-hoc [21,23]. Each feature was individually randomised. The model’s AUC-ROC on the validation sets was then compared to the AUC-ROC before the feature had been randomised. PFI provides an estimate of the extent to which a model relies on a feature for its predictive performance and generalisability. The changes in performance were normalised by the sum of absolute changes over all features. Averages and standard deviations over the validation sets have been reported.

Accumulated local effects (ALE) were computed to determine the directionality of a feature’s effect on model predictions [24]. Specifically, the feature space was divided into ten percentile bins and each feature’s effect was calculated as the difference in predictions between the upper and lower bounds of each bin, leaving all other features unchanged. Binning features in this way can reduce the influence of correlated features often encountered when trying to isolate the effect of a single feature.

## Results

### Patient pathways

A summary of observed patient in-hospital pathways is shown in Figure 1A. Of the 820 patients in cohort A, which we present as an example, 818 (99.8%) entered the hospital via the ED, while 1 (0.1 %) and 1 (0.1%) patients were admitted directly to a ward and the AICU, respectively. Upon leaving the ED, 775 (94.5%) patients transitioned to regular wards and 44 (5.4%) to an AICU. Of the 775 patients in regular wards, 81 (10.5%) patients required subsequent admission to an AICU, 441 (57%) were discharged, 113 (14.5%) remained in hospital and 138 (18%) succumbed to the infection. From the 126 patients that have been admitted to an AICU, 57 (37%) were ultimately discharged, 32 (35%) did not survive and 37 (29%) are still in hospital. Patients’ median length of stay in ED was 5 hours (IQR 3.45 hours). During this time, demographic information, vitals and laboratory values were collected (Fig. 1B). To aid an early patient stratification, our models use data collected during the ED stay only to predict whether a patient reached any of three clinical endpoints during their subsequent admission.

**Figure 1.**
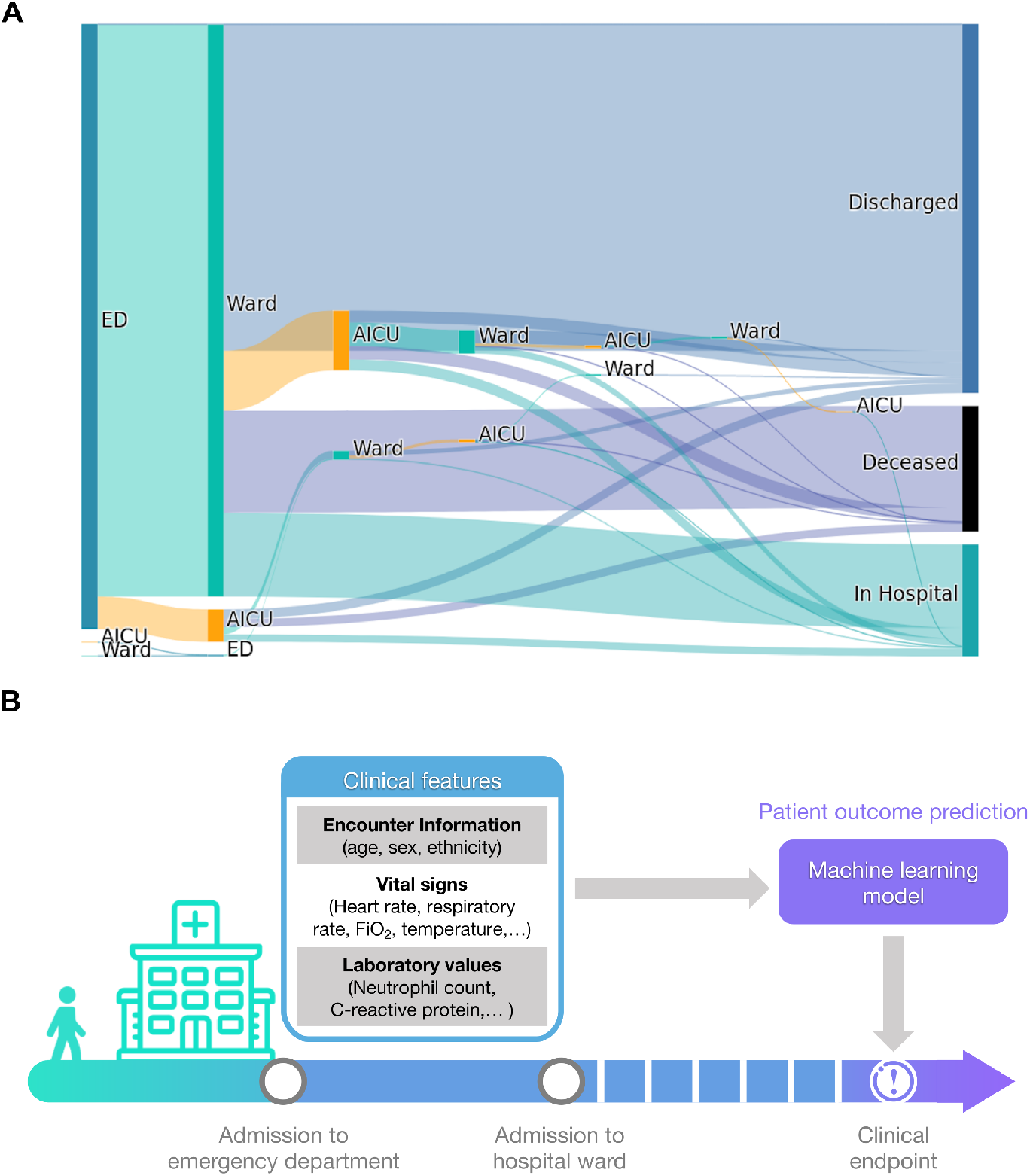
Patient pathways and outcome prediction. **(A)** Patient transitions between hospital departments are shown as bands proportional in size to patient numbers. Different departments are indicted by rectangles (ED, emergency department; Ward, regular hospital ward; AICU, adult intensive care unit). Patients who remain in hospital, are being discharged or die in hospital are indicated on the right. **(B)** Patient outcome prediction models use clinical data recorded within the ED stay of a patient to predict clinical endpoints during the remainder of the in-hospital stay.

### AICU admission

First, we studied patients transitioning to critical care and requiring admission to an AICU. All three models reach good prediction performance on this endpoint, as measured by area under the curve (AUC) of the receiver operating characteristic (ROC) and precision-recall curves, significantly outperforming random classifiers (Fig. 2). The best performing model, XGBoost, reaches an AUC-ROC of 0.83 and an F1 score of 0.51. Both tree-based methods perform better than logistic regression (Table 4). This is to be expected since logistic regression cannot model interactions between features unless such interactions are explicitly encoded into the training data set through feature engineering. All models show a moderate amount of variability across cross-validation folds (notice standard deviations in Fig. 2 and Table 4), which can compromise subsequent analyses. This instability originates from the limited number of patients and high class imbalance between target and control patients (see Table 2). Specifically, in each of the three cross-validation folds the models are only trained and validated on two thirds and one third of the data set, respectively, leaving few target patients for these tasks.

**Figure 2.**
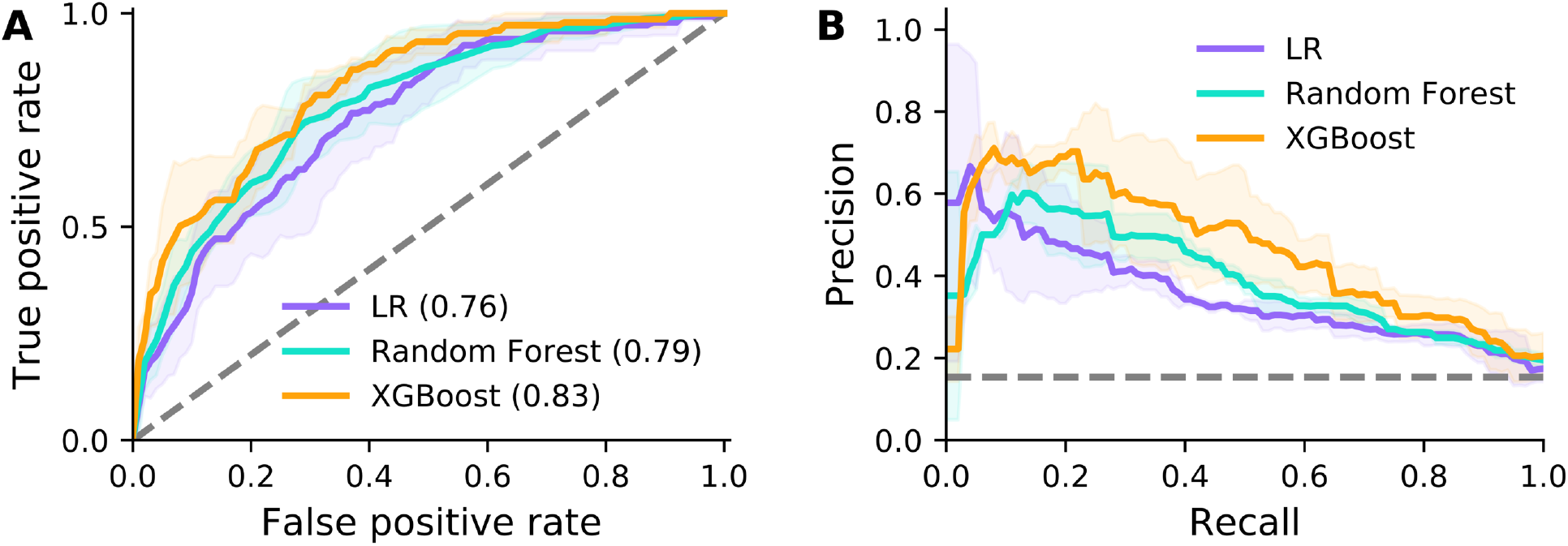
Prediction performance for AICU admission. Model performance for the logistic regression (LR), random forest and XGBoost models are shown as ROC (A) and precision-recall curves (B). AUC under ROC is provided in brackets. Solid lines and shaded areas indicate the mean and standard deviation across three cross-validation folds, respectively. Dashed lines indicate random classifiers.

**Table 4.** Model performance on clinical endpoint prediction (standard deviation shown in brackets).

| Model | Endpoint A<br>(AICU admission) |  | Endpoint B<br>(ventilation) |  | Endpoint C<br>(mortality) |  |
| --- | --- | --- | --- | --- | --- | --- |
|  | AUC | F1 | AUC | F1 | AUC | F1 |
| Logistic regression | 0.76<br>(0.067) | 0.40<br>(0.029) | 0.79<br>(0.097) | 0.41<br>(0.083) | 0.66<br>(0.030) | 0.50<br>(0.035) |
| Random forest | 0.79<br>(0.058) | 0.41<br>(0.031) | 0.81<br>(0.045) | 0.37<br>(0.081) | **0.75**<br>(0.016) | 0.55<br>(0.039) |
| XGBoost | **0.83**<br>(0.045) | **0.51**<br>(0.037) | **0.83**<br>(0.083) | **0.41**<br>(0.052) | 0.74<br>(0.011) | **0.56**<br>(0.035) |

Next, we assessed which clinical variables contribute the most to model predictions by applying PFI. Figure 3A presents the 15 most important features for the logistic regression with elastic net regularisation. Note that clinical variables that can be recorded multiple times during a patient’s ED visit were aggregated to retain only the minimum, maximum, mean and last observation value during the ED stay. Patient age, C-reactive protein and sex reached high importance and significance over cross-validation folds for the logistic regression. Moreover, the fraction of inspired oxygen (FiO_2_) contributes to predictions, albeit without being significant. The random forest (Fig. 3B) and XGBoost (Fig. 3C) models assign a higher importance to patient age, with respiratory rate following thereafter. Intriguingly, ALE analyses reveal that lower patient age increases the likelihood of AICU admission in all three models (Figs. 3D-F). This agrees well with a bias towards younger patients when comparing AICU-admitted patients with control patients (Fig. S3A). However, clinical indicators of disease severity, such as C-reactive protein and ferritin levels, show no clear trend across age groups (Fig. S4). We also find that the fraction of inspired oxygen (Fig. 3D) and respiratory rate (Figs. 3E and F) exhibit a positive effect on AICU admission probability.

**Figure 3.**
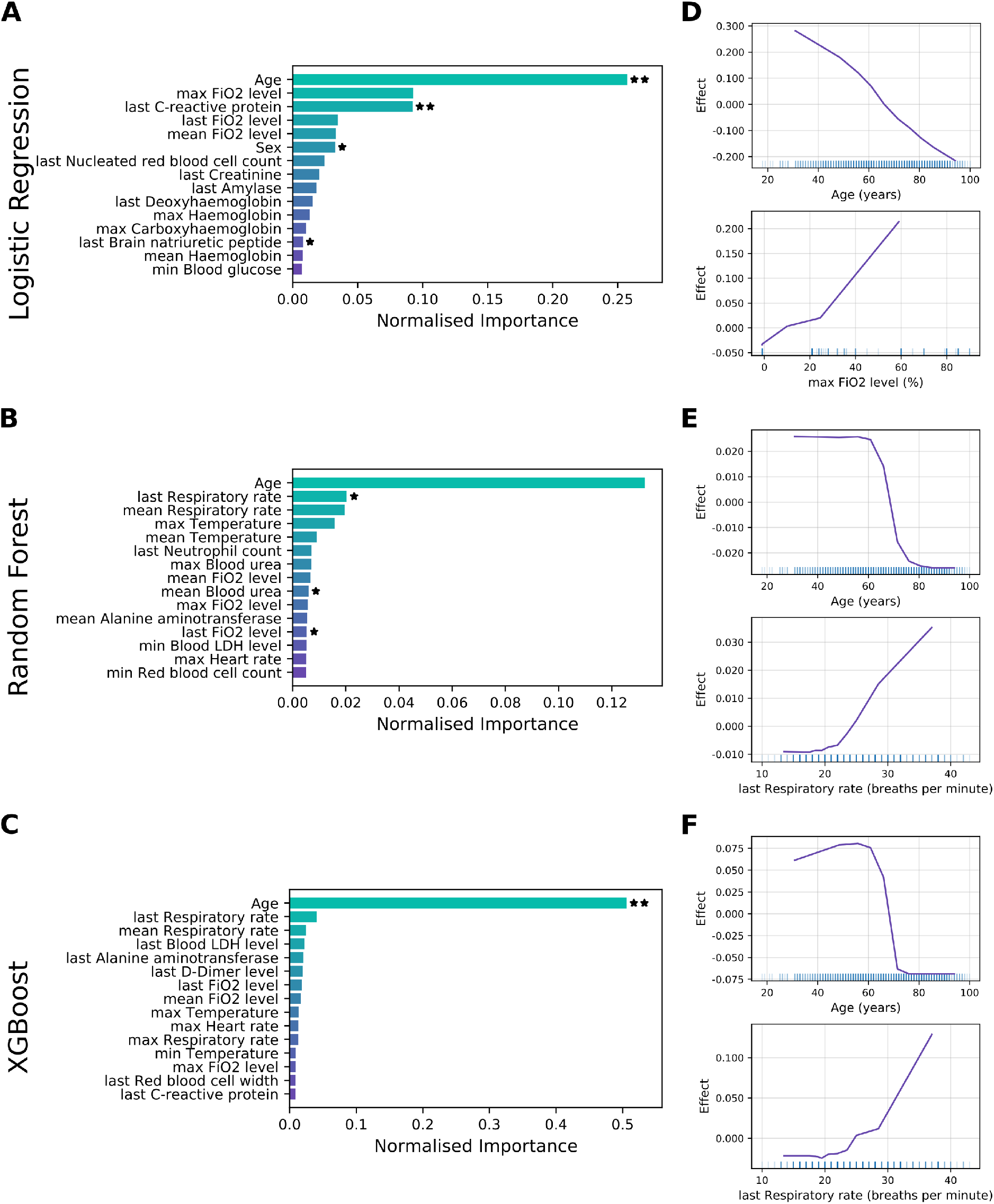
Feature importance for AICU admission. **(A-C)** Permutation feature importance for the logistic regression (A), random forest (B) and XGBoost (C) models. Only the top 15 features are shown. Asterisks mark features with importance scores significantly different from zero across three cross-validation folds with t-test p-value thresholds of 5% (*)and 1% (**). (**D-F**) Accumulated local effects plots for the logistic regression (D), random forest (E) and XGBoost models (F). The top two features according to permutation feature importance are shown for each model. Vertical bars at the bottom indicate feature values observed in the data set.

In summary, machine learning algorithms can predict those patients most likely to require AICU admission in COVID-19 patients from EHR data available during the initial ED stay with high precision. Patient age and indicators of oxygenation status are strong indicators of patient outcome, with advanced age decreasing the probability of AICU admission.

### Mechanical ventilation

For mechanical ventilation prediction, we categorised patients into those that needed a ventilator (e.g., patients receiving SIMV, BIPAP or APRV ventilation) and control patients that either were able to breathe normally or required minimal assistance (e.g., those patients receiving oxygen via nasal cannulae or face masks). Prediction performance on this endpoint is comparable to prediction of AICU admission (Fig. 4). Specifically, XGBoost performs best, reaching an AUC of 0.83, while logistic regression and random forest reach 0.79 and 0.81, respectively (Table 4). This result is expected since most patients receive mechanical ventilation in AICU, meaning the ventilation cohort is a subset of the critical care cohort (56 of 62 target patients in Cohort B are target patients in Cohort A). Notably, all models show a decrease in stability in predicting this clinical endpoint. This is most likely due to a higher class-imbalance and lower number of patients receiving ventilation.

**Figure 4.**
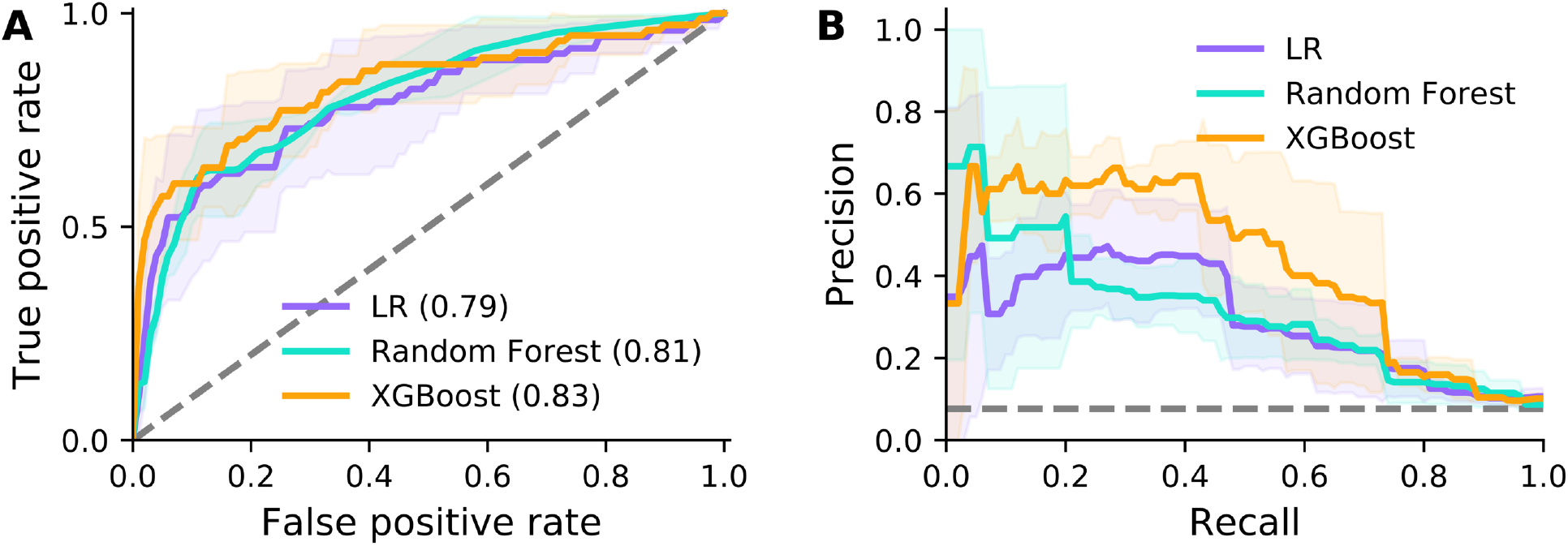
Prediction performance for mechanical ventilation. Model performance for the logistic regression (LR), random forest and XGBoost models are shown as ROC (A) and precision-recall curves (B). AUC under ROC is provided in brackets. Solid lines and shaded areas indicate the mean and standard deviation across three cross-validation folds, respectively. Dashed lines indicate random classifiers.

Feature importance analysis for the logistic regression shows a large effect of the fraction of inspired oxygen and patient age (Fig. 5A). This mirrors the results for AICU admission. We also observe a significant influence of haemoglobin levels on model predictions. Both tree-based methods rank age highly (Figs. 5B and C). In addition, blood lactate levels and oxygen saturation are used by the random forest (Fig. 5B), while XGBoost relies on the fraction of inspired oxygen and levels of thyroid stimulating hormone (Fig. 5C), although few values are significant. In general, all models rely on a broader set of features for the ventilation endpoint. ALE analysis shows younger patients had an increased probability of receiving ventilation (Fig. 5D-F), which agrees with an inherent bias towards younger age when comparing ventilated with non-ventilated patients (Fig.S4B). By contrast, a higher fraction of inspired oxygen and higher blood lactate level were associated with a poor prognosis.

**Figure 5.**
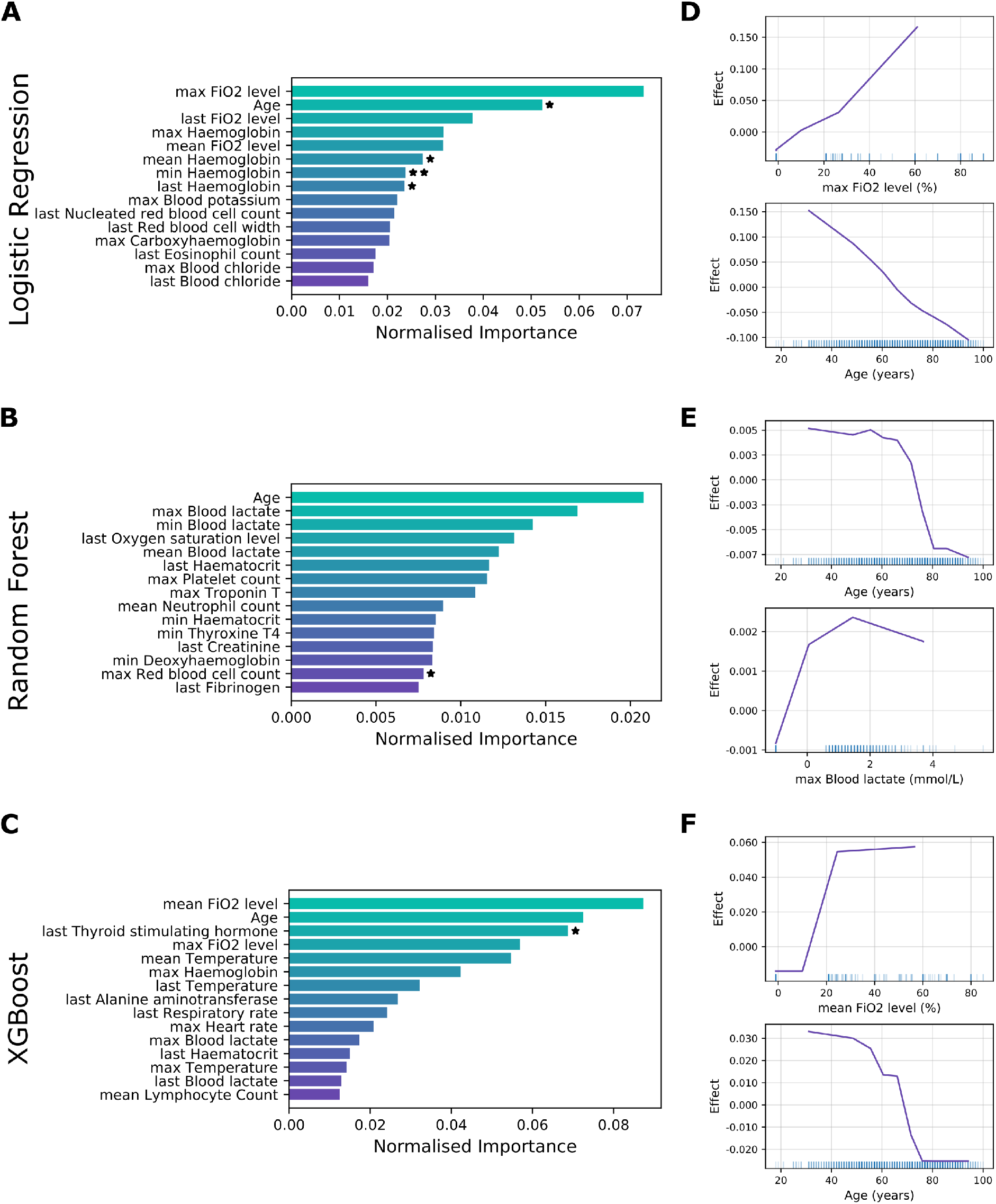
Feature importance for mechanical ventilation. Permutation feature importance for the random forest (A), logistic regression (B) and XGBoost (C) models. Only the top 15 features are shown. Asterisks mark features with importance scores significantly different from zero across three cross-validation folds with t-test p-value thresholds of 5% (*)and 1 % (**). (**D-F**) Accumulated local effects plots for the logistic regression (D), random forest (E) and XGBoost models (F). The top two features according to permutation feature importance are shown for each model. Vertical bars at the bottom indicate feature values observed in the data set.

Taken together, models show good performance when predicting ventilation, albeit with a decreased model stability (higher standard deviation). Patient age and oxygenation status are most predictive of poor outcome, with additional contributions from blood test values, such as lactate and haemoglobin levels.

### Mortality

The performance of all three models shows a marked decrease when predicting mortality (Fig. 6). The logistic regression and XGBoost reach AUCs of 0.66 and 0.74, respectively, only outperformed by random forest reaching an AUC of 0.75. However, model stability is improved with standard deviations across cross-validation folds reaching their lowest levels over all three clinical endpoints (Table 4).

**Figure 6.**
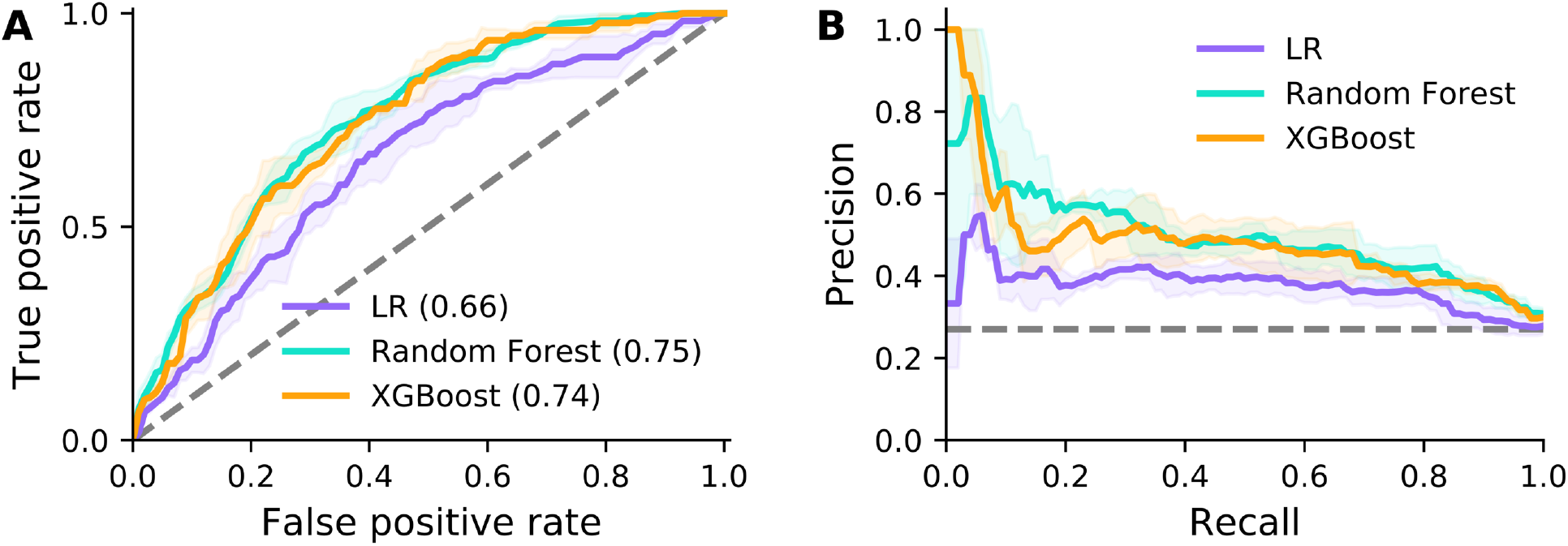
Prediction performance for mortality. Model performance for the logistic regression (LR), random forest and XGBoost models are shown as ROC (A) and precision-recall curves (B). AUC under ROC is provided in brackets. Solid lines and shaded areas indicate the mean and standard deviation across three cross-validation folds, respectively. Dashed lines indicate random classifiers.

Predictions from the logistic regression model are dominated by patient age, with C-reactive protein levels adding a small but significant contribution (Fig. 7A). Similarly, tree-based methods rely heavily on age for their predictions, with smaller contributions of respiratory rate and Troponin T levels (Figs. 7B and C). More generally, prediction of mortality relies more strongly on blood tests as opposed to indicators of oxygen supply observed in other cohorts. ALE analysis shows that advanced age is predictive of higher mortality (Fig. 7D-F). This agrees with a bias towards older age in patients that die in hospital (Fig. S4C). Higher C-reactive protein, respiratory rate and Troponin T levels increase the risk of mortality in our models (Figs. 7D-F).

**Figure 7.**
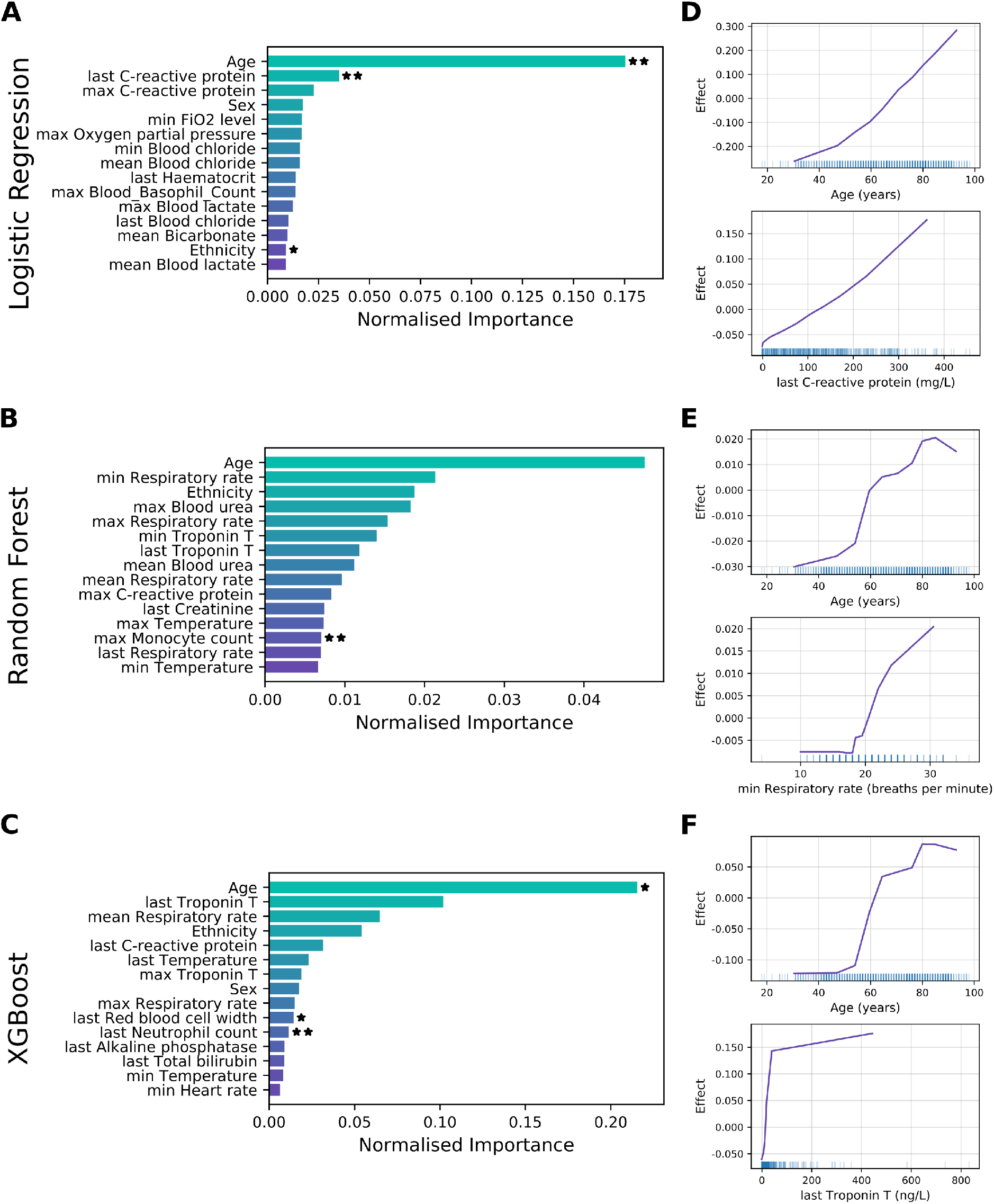
Feature importance for mortality. **(A-C)** Permutation feature importance for the logistic regression (A), random forest (B) and XGBoost (C) models. Only the top 15 features are shown. Asterisks mark features with importance scores significantly different from zero across three cross-validation folds with t-test p-value thresholds of 5% (*)and 1 % (**). (**D-F**) Accumulated local effects plots for the logistic regression (D), random forest (E) and XGBoost models (F). The top two features according to permutation feature importance are shown for each model. Vertical bars at the bottom indicate feature values observed in the data set.

In summary, models show an increased stability but lower overall performance when predicting mortality. Feature importance scores reveal a high and significant contribution of patient age with advanced age contributing to poor patient outcomes.

## Discussion

Disease severity can vary dramatically between COVID-19 patients, ranging from asymptomatic infection to severe respiratory distress and failure. To evaluate the potential of an early stratification of hospitalised patients into risk groups, we built machine learning models from EHR care data of confirmed Covid-19 positive patients, aimed at predicting one of three clinical endpoints: admission to AICU, the need for mechanical ventilation and mortality. On all three cohorts, our models reach good performance with the best model showing AUC-ROC between 0.75 and 0.83. Overall, mortality proved to be the most difficult prediction task, presumably reflecting the complex interactions underlying in-hospital death.

The most predictive feature for all three endpoints was patient age, followed by indicators of patients’ oxygenation status, including fraction of inspired oxygen and respiratory rate. Given that SARS-CoV-2 causes an infection of the respiratory tract, which can lead to severe respiratory distress, these results were to be expected. Our findings are supported by similar works, in which age is consistently found to be the most important feature [16–18]. However, we note that other potential indicators for severe viral infection, like increased temperature and markers of immune system activation, e.g. C-reactive protein, are less prominent in our feature importance scores. Overall, prediction of mortality relies more strongly on blood tests as opposed to indicators of oxygen supply observed in other cohorts. The reason for this observation and its clinical significance is, as of yet, unclear. Our ALE analysis reveals that lower patient age contributes to an increased probability of receiving mechanical ventilation and critical care in AICU, while coinciding with lower mortality. We also note that Docherty *et al*. find that 17% of COVID-19 patients require admission to a High Dependency or Intensive Care Unit [25], which is similar to 15% of patients in our data.

Conversely, our findings concerning the importance of features relating to patients’ oxygenation status are not corroborated by other works. Specifically, other studies find that one important predictor of patient outcome is the level of lactate dehydrogenase [17,18], which, although present in our data set, does not significantly contribute to predictions.

A novel aspect of the present analysis is the use of data limited to a patient’s first few hours in ED. While this perhaps more accurately reflects the data available at the time of admission, it may well come at the cost of missing important information, such as medical history or primary care data, for predicting patient outcome. This may explain the comparative difficulty in predicting mortality, since a patient’s overall chance of surviving infection may depend heavily on their medical history. Also note that, in our analysis, all patients were considered together for mortality prediction and the cohort was not further split according to confounding factors such as age or sex. In addition, mortality data for recent hospital admissions are by their nature censored, with clinical endpoints for patients who remain in hospital not yet fully known.

While we base our study on a comparatively large data set from two hospitals, longitudinal information from additional treatment centres and geographic regions may improve a model’s ability to generalise. We note that such data is currently unavailable for COVID-19. However, future studies may benefit from a multicentre approach. As a result of limited data and the imbalanced cohorts, model stability remains a major challenge. While we use inverse class weights and stratified 3-fold cross validation to mitigate this issue, large uncertainties in model results persist, and many predictions do not reach statistical significance. Increased patient numbers, in particular among target patients, may lead to more conclusive results. Once such data is available, more complex models, such as deep neural networks, may achieve higher prediction performance. A key aspect which should be considered in such works is the prediction horizon, which impacts on how useful a model could be.

In conclusion, our models represent a first step towards the prediction of COVID-19 patient pathways in hospital at the point of admission in the emergency department. While they succeed in predicting patient outcomes and reveal critical clinical variables that may influence patient trajectories, larger data sets and further analyses are required to draw clinically relevant conclusions.

## Data Availability

The data are not publicly available.

## Acknowledgments

This work uses data provided by patients and collected by the NHS as part of their care and support. We believe using patient data is vital to improve health and care for everyone and would, thus, like to thank all those involved for their contribution.

Special thanks are due to the Chelsea and Westminster (CW) COVID-19 AICU Consortium, comprising all critical care personnel who were part of the delivery of care during the COVID-19 pandemic as follows: CW Anaesthetics Consultants, Critical Care Consultants, Trainees & Fellows from ICU, Anaesthesia, and seconded to ICU from other specialities, Surgeons, the supporting Respiratory and ED Physicians, Operating Department Practitioners and CW Critical Care Nurses.This united approach to an unprecedented clinical condition was critical not only to the management of the patients but also to our ability to document and collate the key data in a timely manner to support this analysis.

Also a special thank you to Trystan Hawkin, Chris Chaney from CWplus, the Planned Care Clinical Division managers, porters, domestic personnel and the CW local community who without hesitation have supported the National Healthcare System throughout the COVID-19 pandemic.

## Ethics statement

The data were extracted, anonymised, and supplied by the Trust in accordance with internal information governance review, NHS Trust information governance approval, and General Data Protection Regulation (GDPR) procedures outlined under the Strategic Research Agreement (SRA) and relative Data Sharing Agreements (DSAs) signed by the Trust and Sensyne Health plc on 25th July 2018.

